# SARS-CoV-2 Sequence Characteristics of COVID-19 Persistence and Reinfection

**DOI:** 10.1101/2021.03.02.21252750

**Authors:** Manish C. Choudhary, Charles R. Crain, Xueting Qiu, William Hanage, Jonathan Z. Li

## Abstract

**Background:** Both SARS-CoV-2 reinfection and persistent infection have been reported, but sequence characteristics in these scenarios have not been described. We assessed published cases of SARS-CoV-2 reinfection and persistence, characterizing the hallmarks of reinfecting sequences and the rate of viral evolution in persistent infection.

**Methods:** A systematic review of PubMed was conducted to identify cases of SARS-CoV-2 reinfection and persistence with available sequences. Nucleotide and amino acid changes in the reinfecting sequence were compared to both the initial and contemporaneous community variants. Time-measured phylogenetic reconstruction was performed to compare intra-host viral evolution in persistent SARS-CoV-2 to community-driven evolution.

**Results:** Twenty reinfection and nine persistent infection cases were identified. Reports of reinfection cases spanned a broad distribution of ages, baseline health status, reinfection severity, and occurred as early as 1.5 months or >8 months after the initial infection. The reinfecting viral sequences had a median of 17.5 nucleotide changes with enrichment in the ORF8 and N genes. The number of changes did not differ by the severity of reinfection and reinfecting variants were similar to the contemporaneous sequences circulating in the community. Patients with persistent COVID-19 demonstrated more rapid accumulation of sequence changes than seen with community-driven evolution with continued evolution during convalescent plasma or monoclonal antibody treatment.

**Conclusions:** Reinfecting SARS-CoV-2 viral genomes largely mirror contemporaneous circulating sequences in that geographic region, while persistent COVID-19 has been largely described in immunosuppressed individuals and is associated with accelerated viral evolution.

**Summary:** Reinfecting SARS-CoV-2 viral genomes largely mirror contemporaneous circulating sequences in that geographic region, while persistent COVID-19 has been largely described in immunosuppressed individuals and is associated with accelerated viral evolution.

**Funding:** This study was funded in part by the NIH grant 106701.

**Disclosures:** Dr. Li has consulted for Abbvie.

## BACKGROUND

After resolution of coronavirus disease 2019 (COVID-19) following SARS-CoV-2 infection, antibodies against SARS-CoV-2 persist in the majority of patients for 6 months or more [1]. Despite this, there have now been a number of reports of COVID-19 reinfection, spanning a broad range of age groups, time frames, and disease severity [2-7]. There remains a great deal of uncertainty over the viral characteristics of reinfection cases, including the degree of sequence heterogeneity and the location of new mutations between the initial and reinfecting variants, if any. In addition, the diagnosis of COVID-19 reinfection has been complicated by the increasing reports of persistent COVID-19 infection, especially in immunosuppressed individuals. Like reinfection cases, persistent COVID-19 can also span the range of disease severity, from asymptomatic to severe disease, and recurrent symptoms can last for months [8-11]. Differentiating between persistence and reinfection can be challenging, and little is known about differences in the location and quantity of SARS-CoV-2 mutations in these scenarios. We performed an analysis of SARS-CoV-2 sequences from published cases of COVID-19 reinfection and persistence, characterizing the hallmarks of reinfecting sequences and the rate of viral evolution in persistent infection.

## METHODS

### Data search and selection criteria

We conducted a systematic literature review in PubMed through March 8, 2021 for cases of persistent COVID-19 using the search term “((covid or sars-CoV-2) AND (persistent or persistence or prolonged)) AND (sequence or evolution)”. A search for COVID-19 reinfection reports was made using the terms “(covid or sars-CoV-2) AND (reinfection)”. Both peer-reviewed and preprint results were evaluated. We used the Preferred Reporting Items for Systematic Reviews and Meta-Analyses (PRISMA) for reviewing literature and for reporting search results. Additional preprints that appeared through Google search and that met our criteria were also included. For cases of reinfection, papers were included if the authors described it as a case of reinfection diagnosed >30 days after the initial infection and if whole genome SARS-CoV-2 sequences or sites of mutations relative to a reference sequence (e.g., Wuhan-Hu-1) from both infection time-points were available. Of the 291 results from the search, 14 articles met the inclusion criteria and were included in the present report along with 2 additional preprints that were identified (Supplemental Figure 1A).

**Figure 1.**
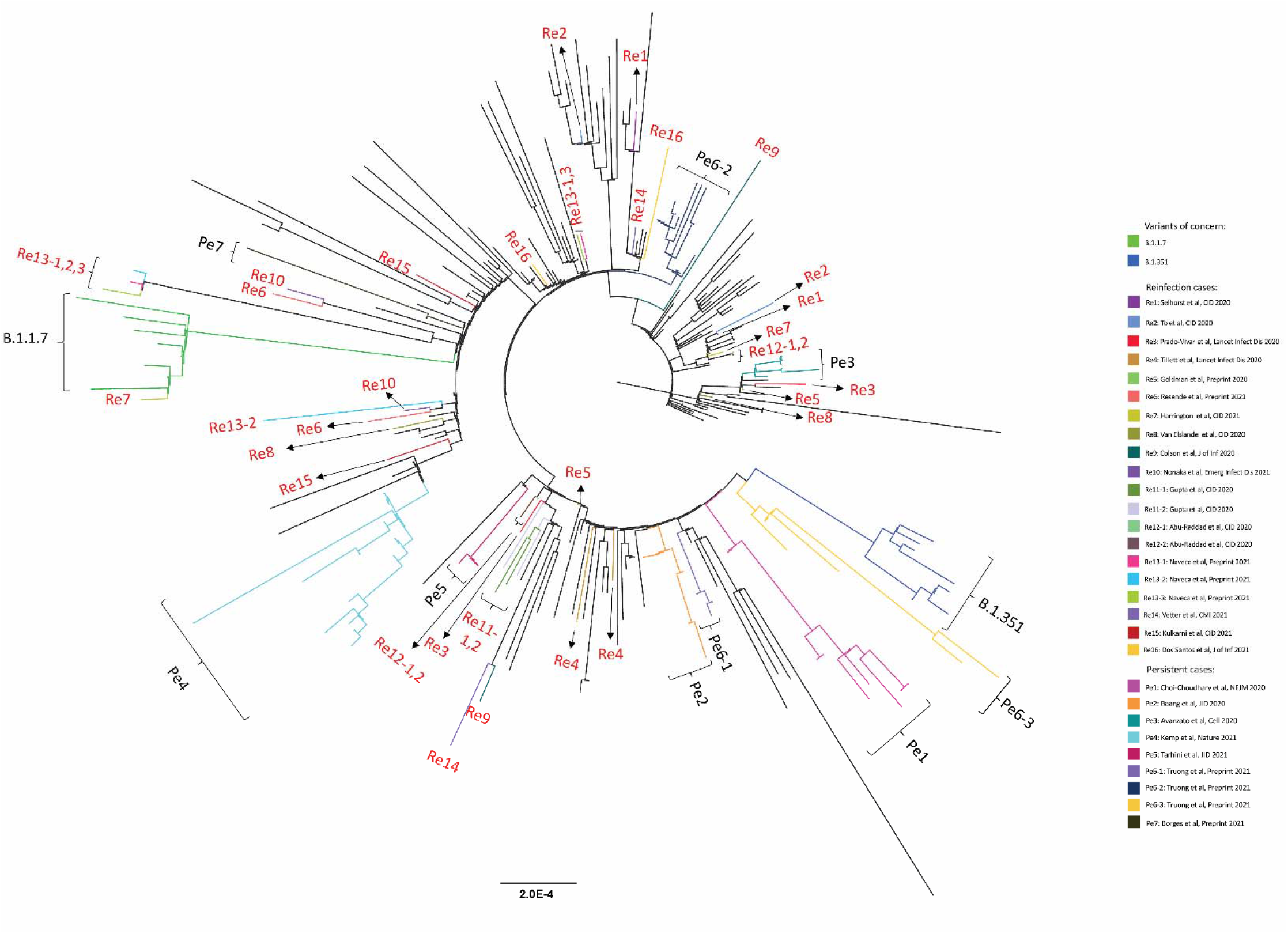
Maximum-likelihood phylogenetic tree of sequences from persistent COVID-19 cases (Pe1-Pe7), COVID-19 reinfection cases (Re1-Re16), the variants of concern B.1.1.7 and B.1.351, and globally sampled sequences from GISAID.

Persistent cases were included if the authors described it as a case of persistent COVID-19 infection and if longitudinal whole genome SARS-CoV-2 sequences were available. The search returned 129 results, 7 of which met the inclusion criteria and were included in the present report along with one other preprint (Supplemental Figure 1B). Only sequences obtained directly from patient respiratory tract samples were included in our analysis to exclude the possibility of sequence changes during the *ex vivo* culture process. Three cases were excluded due to uncertainty in their classification as either reinfection or persistent infection cases (Supplemental Methods, Supplemental Table 1, Supplemental Figure 2).

**Table 1.** Reinfection cases.

| Patient | Authors | Publication/P re-print server (year) | Age | Sex | Comorbidities | Time between infections (days) | Second infection severity | First infection clade | Second infection clade |
| --- | --- | --- | --- | --- | --- | --- | --- | --- | --- |
| Re1 | Selhorst, *et al.* | *Clin. Infect. Dis.* (2020) | 39 | F | None | 185 | Less | 19A | 20A |
| Re2 | To, *et al.* | *Clin. Infect. Dis.* (2020) | 33 | M | None | 144 | Less | 19A | 20E |
| Re3 | Prado-Vivar, *et al.* | *Lancet Infect. Dis.* (2021) | 46 | M | None | 63 | More | 20A | 19B |
| Re4 | Tillett, *et al.* | *Lancet Infect. Dis.* (2020) | 25 | M | None | 48 | More | 20C | 20C |
| Re5 | Goldman, *et al.* | medRxiv | 60-69 | N/A | Emphysema, hypertension | 139 | Less | 19B | 20A |
| Re6 | Resende, *et al.* | Virological | 37 | F | None | 116 | Similar | 20B | 20B |
| Re7 | Harrington, *et al.* | *Clin. Infect. Dis.* (2021) | 78 | M | Diabetic nephropathy with hemodialysis, COPD, sleep apnea, ischemic heart disease | 250 | More | 19A | 20I (B.1.1.7) |
| Re8 | Van Elslande, *et al.* | *Clin. Infect. Dis.* (2021) | 51 | F | Asthma | 93 | Less | 20B | 19B |
| Re9 | Colson, *et al.* | *J. Infect.* (2020) | 70 | M | None | 105 | Less | 20A | 20A.EU2 |
| Re10 | Nonaka, *et al.* | *Emerg. Microbes Infec.* (2021) | 45 | F | None | 147 | More | 20B | 20B |
| Re11-1 | Gupta, *et al.* | *Clin. Infect. Dis* (2020) | 25 | M | None | 108 | Both asymptomatic | 19A | 20A |
| Re11-2 |  |  | 28 | F | None | 111 | Both asymptomatic | 20A | 20A |
| Re12-1 | Abu-Raddad, *et al.* | *Clin. Infect. Dis* (2020) | 25-29 | M | N/A | 46 | N/A | 19A | 20A |
| Re12-2 |  |  | 40-44 | M | N/A | 71 | N/A | 19A | 20A |
| Re13-1 | Naveca, *et al.* | Research Square | 29 | N/A | N/A | 281 | Similar | 20A | 20J (P.1) |
| Re13-2 |  |  | 50 | N/A | N/A | 153 | Similar | 20B | 20J (P.1) |
| Re13-3 |  |  | 40 | F | N/A | 282 | Similar | 20A | 20J (P.1) |
| Re14 | Vetter, *et al.* | *Clin. Microbiol. Infec.* (2021) | 36 | F | None | 205 | Similar | 20A | 20A.EU2 |
| Re15 | Kulkarni, *et al.* | *Clin. Infect. Dis* (2021) | 61 | M | None | 44 | More | 20B | 20B |
| Re16 | Adrielle dos Santos, *et al.* | *J. Infect.* (2021) | 40 | F | Systemic arterial hypertension, obesity | 53 | Less severe | 20A | 20A |

**Figure 2.**
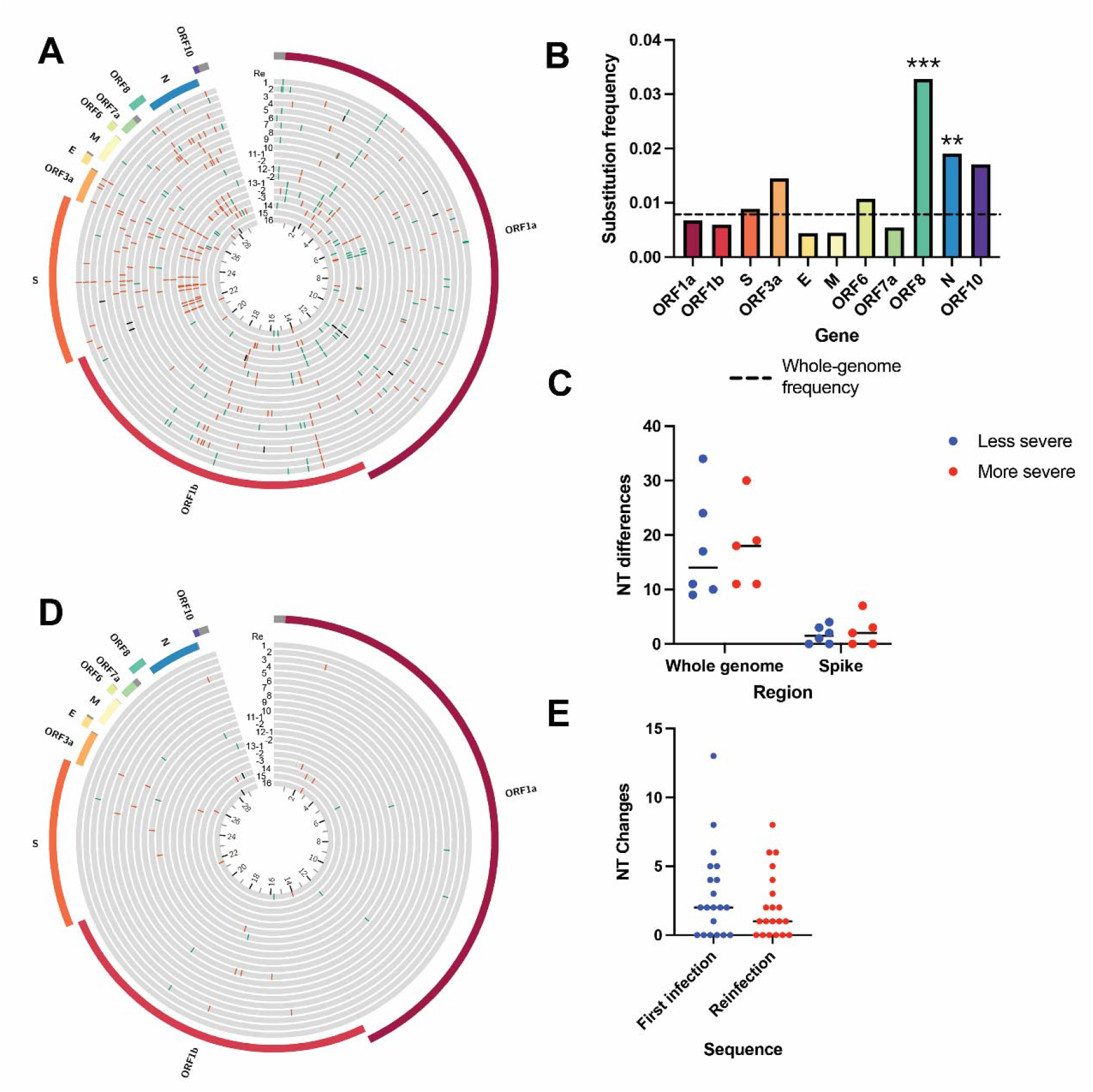
Comparison of viral sequences from reinfection cases. (A) Circos plot showing location of nucleotide changes in the reinfecting sequence relative to the initial infection sequence for each of the 20 cases. Inner ring indicates nucleotide position in kilobases. Synonymous changes are in green, nonsynonymous changes in orange, deletions in black. (B) Nucleotide (NT) substitution frequency pooled across all reinfection cases for each SARS-CoV-2 gene. Dashed line indicates global substitution frequency across the whole genome. Substitution frequency for each gene was compared to the substitution frequency in the rest of the genome using a Fisher’s exact test. P-values were corrected for multiple comparisons using the Bonferroni correction. * <0.05, ** <0.01 and ***<0.001 (C) Nucleotide changes in the second infection relative to the first infection by clinical disease severity. Mutations shown for the whole genome and S gene. P=0.67, Mann Whitney test. (D) Circos plot showing location of nucleotide mutations from the second infection relative to other viruses circulating at the same time in the same geographic region. Only rare mutations present in <1% of contemporaneous community sequences are shown. (E) Number of rare nucleotide polymorphisms at each time point relative to circulating sequences in the community. P=0.26, Wilcoxon matched-pairs signed rank test. ORF: open reading frame, S: Spike, E: Envelope, M: Membrane, N: Nucleocapsid.

Sequences were analyzed for mutations using NextClade (https://clades.nextstrain.org/) and snp-sites (https://github.com/sanger-pathogens/snp-sites). The degree of reinfection severity, either more or less severe compared to the first infection, was classified based on an explicit determination by the authors of each article or by comparing symptoms, duration of illness, and hospitalization status between both episodes.

### Sequencing dataset compilation and phylogenetic tree construction

The sequencing dataset contained a total of 262 globally representative SARS-CoV-2 genomes selected from GISAID and sequences from the reinfection and persistence cases (Supplemental Methods; Supplemental Data 1). The sampled sequences were chosen to be representative of global sequence diversity throughout the time course of the pandemic. Sequences of variants of concern B.1.1.7 and B.1.351 were also included. Nucleotide sequence alignment was performed using MAFFT (Multiple Alignment using Fast Fourier Transform) [12]. Best-fit nucleotide substitution was calculated using model selection followed by maximum likelihood (ML) phylogenetic tree construction using IQ-Tree with 1000-bootstrap replicates [12].

### Mutation analysis

For reinfection cases, mutations were determined in two ways. First, nucleotide and amino acid changes were identified for the reinfection sequences relative to the first infection sequence. The frequency of nucleotide or amino acid changes within each gene was compared to the frequency of changes in the remainder of the genome by Fisher’s exact tests with a Bonferroni correction (for multiple comparisons). The relationship between disease severity and number of nucleotide or amino acid changes in the genome was assessed using a Mann-Whitney test. Second, to identify unique characteristics of reinfecting viruses, each of the first and reinfection sequences were compared to circulating sequences in the community as defined by the same NextStrain clade sampled within one month obtained from the same geographic location uploaded to GISAID (https://www.gisaid.org/; Supplemental Table 2, Supplemental Methods, Supplemental Data 2). Rare mutations were determined as polymorphisms that were present only in the reinfecting sequence (not the initial variant) and found in less than 1% of contemporaneous community sequences. Mutation locations are graphically represented in Circos plots [13].

**Table 2.** Persistent cases.

| Patient | Case | Publication (year) | Age | Sex | Underlying conditions | Immunosuppressants | Antiviral treatment | Infection length | Fatal ? |
| --- | --- | --- | --- | --- | --- | --- | --- | --- | --- |
| Pe1 | Choi, *et al.* | *New Engl. J. Med.* (2020) | 45 | M | Antiphospholipid antibody syndrome | Rituximab, ecilizumab, cyclophosphamide, corticosteroids | Remdesivir, casirivimab and imdevimab | 154 | Yes |
| Pe2 | Baang, *et al.* | *J. Infect. Dis.* (2021) | 60 | M | Refractory mantle cell lymphoma | CD20 bispecific Ab, B-cell directed Ab, cyclophosphamide, doxorubicin, prednisone | Remdesivir, convalescent plasma | 156 | Yes |
| Pe3 | Avanzato, *et al.* | *Cell* (2020) | 71 | F | CLL, acquired hypogammaglobulinemia | - | Convalescent plasma | 156 | No |
| Pe4 | Kemp, *et al.* | *Nature* (2021) | - | - | Marginal B cell lymphoma, hypogammaglobulinemia | B-cell depleting therapy | Remdesivir, convalescent plasma | 102 | Yes |
| Pe5 | Tarhini, *et al.* | *J. Infect. Dis.* (2021) | 66 | M | HIV, multifocal leukoencephalopathy from JC virus | - | - | 124 | No |
| Pe6-1 | Truong, *et al.* | medRxiv | <5 | F | B-cell ALL | Dexamethasone, vincristine, peg-asparaginase, methotrexate, 6-MP, doxorubicin | - | 91 | No |
| Pe6-2 |  |  | 20-25 | M | B-cell ALL | CD-19 directed CAR-T cell therapy (tisagenlecleucel), cyclophosphamide, fludarabine | Remdesivir, convalescent plasma | 250+ | No |
| Pe6-3 |  |  | <5 | M | B-cell ALL | Cyclophosphamide,<br>cytarabine, thioguanine,<br>vincristine,<br>dexamethasone,<br>methotrexate,<br>mercaptopurine | - | 196 | No |
| Pe7 | Borges,<br>*et al.* | Virological | 61 | F | Diffuse large B cell<br>lymphoma | Chemotherapy<br>(unspecified),<br>methotrexate,<br>corticosteroids | Remdesivir | 197 | No |

ALL: acute lymphoblastic leukemia, CLL: chronic lymphocytic leukemia, RDV: remdesivir, CP: convalescent plasma, Ab: antibody

For persistent infections, sequence changes were assessed at two time intervals: before or after convalescent plasma or monoclonal antibody treatment. Sequences sampled before convalescent plasma or antibody treatment were compared to the first sequence sampled. For sequences sampled after convalescent plasma or antibody treatment, sequence changes (both nucleotide and amino acid) were determined relative to the last pre-treatment sequence. Linear regression was used to estimate the rate of viral changes between two intervals. The slope of the trendline was compared to the latest global clock rate (March 29, 2021) as estimated by NextStrain (https://nextstrain.org/ncov/global/).

### Time-measured phylogenetic analysis

The temporal signal of the ML tree was examined in TempEst [14] regressing on root-to-tip divergence, and outliers were inspected in the distribution of residuals. A high degree of clock-like behavior in the whole dataset was observed (R^2^ = 0.721) in root-to-tip regression analysis with the slope rate as 8.26E-4 and the rough ancestral time of the sample was calculated as 2019.84. This suggests that the whole dataset has a realistic temporal signal and it is appropriate for an estimation of temporal parameters. No outliers were found in this sample. To further examine the temporal signal in the sequences from persistent patients (especially these with > 2 sequences), separate root-to-tip regression analysis also supported temporal signal for a time-measured phylogeny. To compare the evolutionary rates between the reported persistent infections and the general population infections, time-measured phylogenetic reconstruction was conducted in Bayesian Evolutionary Analysis Sampling Trees (BEAST) v1.10.4 [15]. Nine partitions, including eight persistent patients and the global sequences, were used as separate groups of taxa, to estimate separate evolutionary rates. Due to large uncertainties with small samples, persistent patients with only two viral sequences were excluded from this analysis. A general time reversible (GTR) model was applied with gamma-distributed rate variations among sites. A lognormal relaxed molecular clock was used with an initial mean of 0.0008 and a uniform prior ranging from 0.0 to 1.0. A logistic growth tree prior was applied. Four independent Bayesian Markov Chain Monte Carlo (MCMC) chains of 100 million generations were performed with a sampling step every 10,000 generations to yield 10,000 trees per run. To ensure a sufficient effective sample size ESS > 200, the convergence of three runs was diagnosed in Tracer v 1.7.1 (http://tree.bio.ed.ac.uk/software/tracer/) for all parameters. LogCombiner v1.10.4 as part of the BEAST software package was used to combine the multiple runs to generate log and tree files after appropriate removal of the burn-in from each MCMC chain. The comparison of the evolutionary rates from the combined log file is analyzed and visualized in R v4.0.2 (https://www.r-project.org/).

### Statistical analysis

Nonparametric Wilcoxon rank sum or matched pairs signed rank tests were used to compare the number of amino acid changes between sequences. Statistical analyses were performed using GraphPad Prism 9 (GraphPad Software, San Diego, CA).

## RESULTS

### Sequence analysis of reinfection cases

A total of twenty cases from sixteen reports were included in this analysis (Table 1) [2-7, 16-25]. A broad range of age groups were represented and 90% were under the age of 70 years. Most (80%) of the cases had no reported comorbidities and while one patient had diabetes and end-stage renal disease, none had high-level immunosuppression. The interval between diagnosis of the first infection and the second infection ranged from 44 days to 282 days with a median of 113.5 days. Five patients had more severe illness during the second infection, while six had less severe symptoms on reinfection, including two who were asymptomatic on reinfection. Two cases were asymptomatic in both infections, five cases reported the same severity for both infections and no information on infection severity was available for two cases (Table 1). Six cases reported reinfection with a virus from the same clade.

Phylogenetic analysis demonstrated distinct branching for the two sequences in each of the reinfection cases, corroborating results discussed in the original reports (Figure 1). We compared nucleotide and amino acid changes in the reinfecting viral sequence compared to the initial sequence and found a median of 17.5 nucleotide changes (range 9-37) and 9 amino-acid changes (range 6-24) compared to the original sequence (Figure 2A). The nucleotide changes between the initial and reinfecting sequences were distributed across the SARS-CoV-2 genome, with significantly higher frequencies of changes in ORF8 (P<0.001) and N (P=0.001) (Figure 2B). A similar pattern was observed with amino acid changes (Supplemental Figure 3A). All but two reinfection cases had at least one substitution or deletion in the S gene (Supplemental Table 3). Next, we assessed whether reinfection with a more divergent second virus resulted in more severe disease. We found no significant differences in the number of nucleotide or amino acid changes in the reinfecting virus compared to the original viral variant when categorized by the severity of the reinfection (Figure 2C; Supplemental Figure 3B). Both the initial and reinfecting SARS-CoV-2 variants were similar to the sequences circulating in the community at the time of reinfection. The initial infecting variant harbored a median of only 2 rare nucleotide mutations compared to contemporaneous circulating variants in the community and the reinfecting variant contained a median of only 1 rare nucleotide mutation (Figure 2D-E; Supplemental Figure 3C).

**Figure 3.**
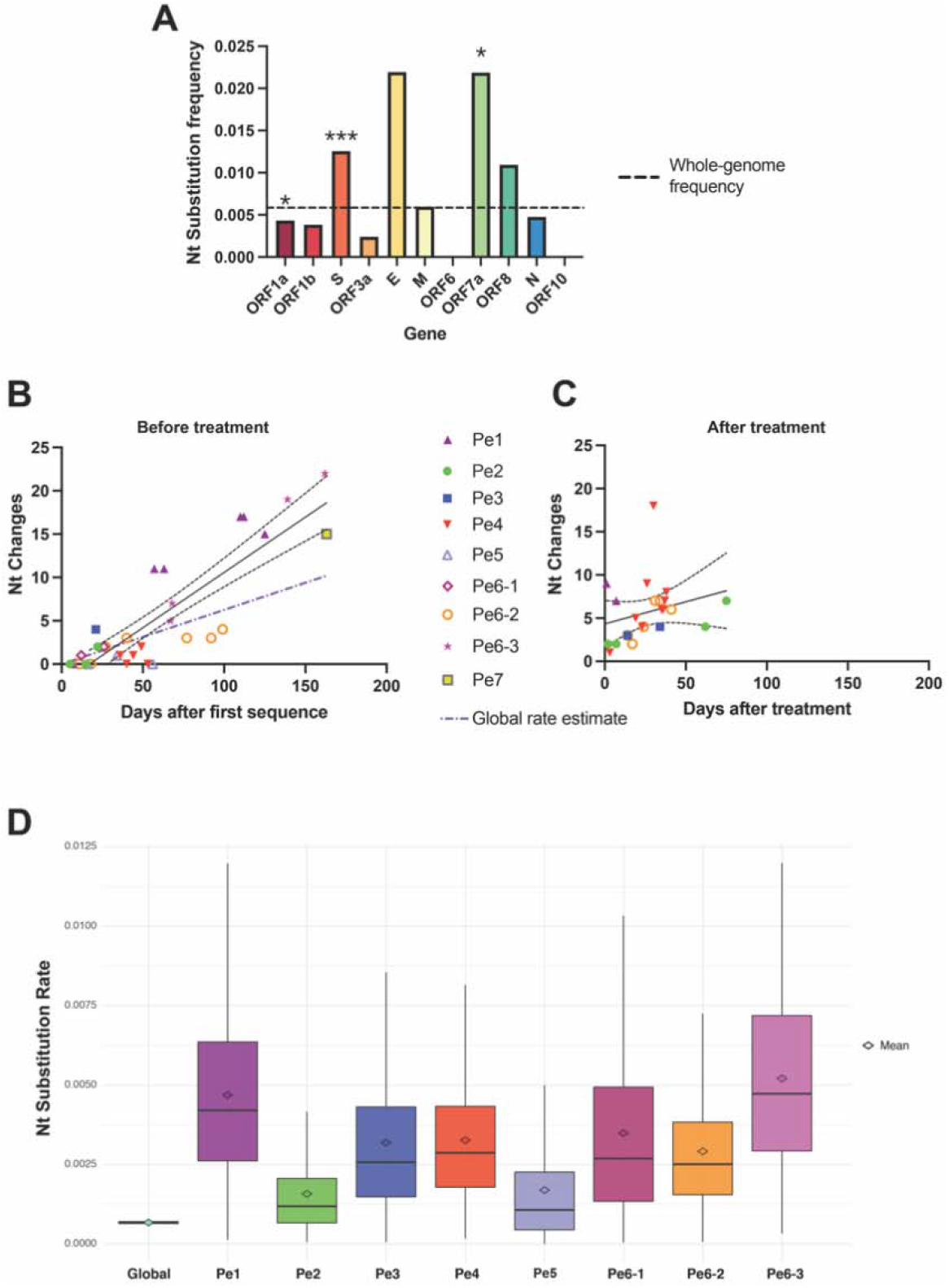
SARS-CoV-2 mutation location and evolutionary rate in the persistent COVID-19 cases. (A) Nucleotide (NT) substitution frequency pooled across all persistent cases for each SARS-CoV-2 gene. Dashed line indicates global substitution frequency across the whole genome. Substitution frequency for each gene was compared to the substitution frequency in the rest of the genome using a Fisher’s exact test. P-values were corrected for multiple comparisons using the Bonferroni correction. * <0.05, ** <0.01 and ***<0.001 (B) Nucleotide changes in samples taken prior to convalescent plasma or monoclonal antibody treatment relative to first sampled sequence in each persistently infected patient. Regression line and 95% confidence bands are shown. Purple dash-dotted line is global rate estimate obtained from NextStrain. (C) Nucleotide changes in samples taken after convalescent plasma or monoclonal antibody treatment relative to last sample taken prior to treatment in each persistently infected patient. Regression line and 95% confidence bands are shown. (D) Substitution rate (nucleotide substitutions per site per year) of sampled global SARS-CoV-2 sequences relative to persistent patients based on Markov chain Monte Carlo time-measured phylogenetic reconstruction. Box plots show median and interquartile ranges of estimated substitution rates. The mean, median, and 95% highest posterior density (HPD) interval can be found in Supplemental Table 4.

### Sequence analysis of persistent COVID-19 cases

A total of nine cases from seven reports describing persistent infection were retrieved from our literature search. Of these nine cases, all but one had B cell immunodeficiency [8-10, 26-29]. Four were treated with B cell-depleting therapy for lymphoma or autoimmune disorders, while four had B cell lymphomas treated with chemotherapy (Table 2). One patient had advanced HIV infection with a CD4+ count of 0 cells/mm^3^ and diminished CD19+ cell counts. The median length of infection was 154 days and 33% of the cases ended in death. One patient had asymptomatic disease throughout [9]. Four patients were treated with convalescent plasma at least once during their illness [9, 10, 26, 28], and one patient was treated with the monoclonal antibodies casirivimab and imdevimab [8].

Phylogenetic analysis revealed that, for each of the nine patients, sequences formed a distinct cluster, confirming what was found in the original reports (Figure 1). New mutations emerging over time were detected in all of the persistent COVID-19 patients with further changes identified after treatment with convalescent plasma or monoclonal antibodies (Supplemental Figure 4). Mutations occurred with significantly higher frequency in S (P<0.001) and ORF7a (P=0.02) and lower frequency in ORF1a (P=0.02) (Figure 3A; Supplemental Figure 5A). The rate of viral evolution was plotted for each patient both for the interval before and after convalescent plasma/antibody treatment. Before antiviral treatment, the rate of sequence changes over time appeared faster than the NextStrain estimate for the global rate of SARS-CoV-2 evolution (dotted purple line) (Figure 3B; Supplemental Figure 5B). Treatment with convalescent plasma or antibody cocktail treatment was insufficient to halt intra-host viral evolution (Figure 3C; Supplemental Figure 5C).

We also performed time-measured phylogenetic reconstruction with the pre-treatment persistent sequences to compare the rate of intra-host viral evolution in persistent COVID-19 to the rate of community-driven evolution. This analysis provided further evidence that SARS-CoV-2 evolution appeared faster in these persistent infection individuals compared to the rate in the general public population, though substantial uncertainties are shown in these estimates given the limited sequence sampling in each patient (Figure 3D; Supplemental Table 4).

## DISCUSSION

We conducted a systematic review and pooled analysis of sequences from reports of COVID-19 reinfection and persistent infection. Reports of reinfection cases demonstrate a wide range of situations: spanning a broad distribution of ages, baseline health status and reinfection severity compared to the initial infection. Reinfection occurred as early as 1.5 months or >8 months after the initial infection. Common explanations for the presence of reinfection involves either waning SARS-CoV-2 antibodies or the presence of viral escape mutations [30, 31]. While most cases of SARS-CoV-2 reinfection did involve infection with a different clade (including the variants of concern B.1.1.7 and P.1), it is noteworthy that mutations were identified throughout the genomes and the frequency of mutations within the S gene was not elevated relative to the rest of the genome. In addition, individuals with more severe reinfections did not have significantly greater frequency of S gene mutations. Interestingly, the genes with the highest frequency of mutations was ORF8 and N. ORF8 is a rapidly evolving accessory protein that may antagonize host immune function [32] while the nucleocapsid is a vital structural protein that also serves as a target for both humoral and cell-mediated immune responses [33]. Finally, the presence of rare mutations was uncommon in the reinfecting virus, which largely mirrored the contemporaneously circulating variants in the region of infection. However, the reinfecting variants generally contained a substantial number of mutations compared to the initial variant, including frequent changes in the S gene, and additional studies are needed to assess whether these changes may have contributed to the risk of repeat infection.

While the number of immunosuppressed individuals with available sequences remains limited, the results suggest that the rate of viral evolution (measuring both synonymous and non-synonymous changes), is accelerated within immunosuppressed individuals. In addition, treatment with convalescent plasma or monoclonal antibody cocktails was insufficient to fully halt viral evolution and the emergence of viral escape with treatment has been documented [26, 34]. Mutations associated with immune escape and/or more efficient replication kinetics, including E484K, S494P, N501Y and N-terminal spike deletions, have been observed in both immunosuppressed individuals and the novel variants of concern [35, 36]. The results raise the possibility that novel variants, including those harboring escape mutations against current treatments, could arise from immunosuppressed individuals and suggest that immunosuppressed individuals should be a focus of public health efforts. Amongst the current reports of persistent COVID-19, B-cell dysfunction appears to be a common thread, including in reports that were not included in this analysis due to a lack of available full-length sequences [37-41]. It is important to note, though, that T cell function may also play a role in protection against SARS-CoV-2 [42] and a subset of these patients also included concurrent suppression of other aspects of the immune response. Additional studies are needed to fully define the type and intensity of immunosuppression that would place patients at greatest risk of persistent COVID-19.

Two factors generally differentiated between reinfection and persistent infection scenarios: first, reinfections have so far been largely described in immunocompetent individuals while the majority of persistent COVID cases have been in immunosuppressed patients. Secondly, phylogenetic analysis can generally differentiate between reinfection and persistent infection, especially in cases where persistent infection allowed the longitudinal collection of >2 sequences. However, given the slow rate of SARS-CoV-2 evolution and limited viral diversity [43], it can be challenging to differentiate between reinfection and persistent infection, especially in situations with limited sampling and/or duration between samples.

A limitation of this work is that it relies on case reports, which can be influenced by publication bias and limits our statistical power. However, to date, there have been no systematic, large-scale sequence-based studies of COVID-19 reinfection or persistent infections. This is partly due to the rarity of these types of cases and that initial infecting sequences are frequently unavailable for comparison with reinfecting or persistently infecting variants. Overall, our results demonstrate the need to further explore factors that increase the risk of breakthrough reinfections and persistent COVID-19. This line of investigation will have important implications on the durability of current available vaccines and for preventing the rise of novel variants.

## Supporting information

Supplemental doc

## Data Availability

NA

## Acknowledgements

We thank Jeremy Luban and Ronald Bosch for their feedback and discussion.

## Notes

### Competing Interest Statement

The authors have declared no competing interest.

### Summary of Updates

Figure 1, 2 & 3 revised; discussion updated.

## REFERENCES

1. Dan JM, Mateus J, Kato Y, et al. Immunological memory to SARS-CoV-2 assessed for up to 8 months after infection. Science 2021.

2. Van Elslande J, Vermeersch P, Vandervoort K, et al. Symptomatic SARS-CoV-2 reinfection by a phylogenetically distinct strain. Clin Infect Dis 2020.

3. To KK, Hung IF, Ip JD, et al. COVID-19 re-infection by a phylogenetically distinct SARS-coronavirus-2 strain confirmed by whole genome sequencing. Clin Infect Dis 2020.

4. Tillett RL, Sevinsky JR, Hartley PD, et al. Genomic evidence for reinfection with SARS-CoV-2: a case study. Lancet Infect Dis 2021; 21(1): 52–8.

5. Selhorst P, Van Ierssel S, Michiels J, et al. Symptomatic SARS-CoV-2 reinfection of a health care worker in a Belgian nosocomial outbreak despite primary neutralizing antibody response. Clin Infect Dis 2020.

6. Harrington D, Kele B, Pereira S, et al. Confirmed Reinfection with SARS-CoV-2 Variant VOC-202012/01. Clin Infect Dis 2021.

7. Goldman JD, Wang K, Roltgen K, et al. Reinfection with SARS-CoV-2 and Failure of Humoral Immunity: a case report. medRxiv 2020.

8. Choi B, Choudhary MC, Regan J, et al. Persistence and Evolution of SARS-CoV-2 in an Immunocompromised Host. N Engl J Med 2020; 383(23): 2291–3.

9. Avanzato VA, Matson MJ, Seifert SN, et al. Case Study: Prolonged Infectious SARS-CoV-2 Shedding from an Asymptomatic Immunocompromised Individual with Cancer. Cell 2020; 183(7): 1901–12 e9.

10. Baang JH, Smith C, Mirabelli C, et al. Prolonged Severe Acute Respiratory Syndrome Coronavirus 2 Replication in an Immunocompromised Patient. J Infect Dis 2021; 223(1): 23–7.

11. Kemp SA, Collier DA, Datir R, et al. Neutralising antibodies in Spike mediated SARS-CoV-2 adaptation. medRxiv 2020.

12. Trifinopoulos J, Nguyen LT, von Haeseler A, Minh BQ. W-IQ-TREE: a fast online phylogenetic tool for maximum likelihood analysis. Nucleic Acids Res 2016; 44(W1): W232–5.

13. Krzywinski MI, Schein JE, Birol I, et al. Circos: An information aesthetic for comparative genomics. Genome Research 2009.

14. Rambaut A, Lam TT, Max Carvalho L, Pybus OG. Exploring the temporal structure of heterochronous sequences using TempEst (formerly Path-O-Gen). Virus Evol 2016; 2(1): vew007.

15. Drummond AJ, Suchard MA, Xie D, Rambaut A. Bayesian phylogenetics with BEAUti and the BEAST 1.7. Mol Biol Evol 2012; 29(8): 1969–73.

16. Prado-Vivar B, Becerra-Wong M, Guadalupe JJ, et al. A case of SARS-CoV-2 reinfection in Ecuador. Lancet Infect Dis 2020.

17. Resende PC, de Vasconcelos RHT, Arantes I, et al. Spike E484K mutation in the first SARS-CoV-2 reinfection case confirmed in Brazil. Virologica 2021.

18. Colson P, Finaud M, Levy N, Lagier JC, Raoult D. Evidence of SARS-CoV-2 re-infection with a different genotype. J Infect 2020.

19. Nonaka CKV, Franco MM, Graf T, et al. Genomic Evidence of a Sars-Cov-2 Reinfection Case With E484K Spike Mutation in Brazil. Preprintsorg 2021.

20. Gupta V, Bhoyar RC, Jain A, et al. Asymptomatic reinfection in two healthcare workers from India with genetically distinct SARS-CoV-2. Clin Infect Dis 2020.

21. Abu-Raddad LJ, Chemaitelly H, Malek JA, et al. Assessment of the risk of SARS-CoV-2 reinfection in an intense re-exposure setting. Clin Infect Dis 2020.

22. Naveca FG, da Costa C, Nascimento V, et al. Three SARS-CoV-2 reinfection cases by the new Variant of Concern (VOC) P.1/501Y.V3. Research Square 2021.

23. Vetter P, Cordey S, Schibler M, et al. Clinical, virologic and immunologic features of a mild case of SARS-CoV-2 reinfection. Clin Microbiol Infect 2021.

24. Kulkarni O, Narreddy S, Zaveri L, Kalal IG, Tallapaka KB, Sowpati DT. Evidence of SARS-CoV-2 reinfection without mutations in Spike protein. Clin Infect Dis 2021.

25. Adrielle Dos Santos L, Filho PGG, Silva AMF, et al. Recurrent COVID-19 including evidence of reinfection and enhanced severity in thirty Brazilian healthcare workers. J Infect 2021; 82(3): 399–406.

26. Kemp SA, Collier DA, Datir RP, et al. SARS-CoV-2 evolution during treatment of chronic infection. Nature 2021.

27. Tarhini H, Recoing A, Bridier-Nahmias A, et al. Long term SARS-CoV-2 infectiousness among three immunocompromised patients: from prolonged viral shedding to SARS-CoV-2 superinfection. J Infect Dis 2021.

28. Truong TT, Ryutov A, Pandey U, et al. Persistent SARS-CoV-2 infection and increasing viral variants in children and young adults with impaired humoral immunity. medRxiv 2021.

29. Borges V, Isidro J, Cunha M, et al. Long-term evolution of SARS-CoV-2 in an immunocompromised patient with non-Hodgkin lymphoma. Virological 2021.

30. Wibmer CK, Ayres F, Hermanus T, et al. SARS-CoV-2 501Y.V2 escapes neutralization by South African COVID-19 donor plasma. bioRxiv 2021.

31. Weisblum Y, Schmidt F, Zhang F, et al. Escape from neutralizing antibodies by SARS-CoV-2 spike protein variants. Elife 2020; 9.

32. Flower TG, Buffalo CZ, Hooy RM, Allaire M, Ren X, Hurley JH. Structure of SARS-CoV-2 ORF8, a rapidly evolving coronavirus protein implicated in immune evasion. bioRxiv 2020.

33. Nelde A, Bilich T, Heitmann JS, et al. SARS-CoV-2-derived peptides define heterologous and COVID-19-induced T cell recognition. Nat Immunol 2021; 22(1): 74–85.

34. Starr TN, Greaney AJ, Addetia A, et al. Prospective mapping of viral mutations that escape antibodies used to treat COVID-19. Science 2021; 371(6531): 850–4.

35. Volz E, Mishra S, Chand M, et al. Assessing transmissibility of SARS-CoV-2 lineage B.1.1.7 in England. Nature 2021.

36. Tegally H, Wilkinson E, Giovanetti M, et al. Detection of a SARS-CoV-2 variant of concern in South Africa. Nature 2021.

37. Camprubi-Ferrer D, Gaya A, Marcos MA, et al. Persistent replication of SARS-CoV-2 in a severely immunocompromised treated with several courses of remdesivir. Int J Infect Dis 2020.

38. Hensley MK, Bain WG, Jacobs J, et al. Intractable COVID-19 and Prolonged SARS-CoV-2 Replication in a CAR-T-cell Therapy Recipient: A Case Study. Clin Infect Dis 2021.

39. Martinot M, Jary A, Fafi-Kremer S, et al. Remdesivir failure with SARS-CoV-2 RNA-dependent RNA-polymerase mutation in a B-cell immunodeficient patient with protracted Covid-19. Clin Infect Dis 2020.

40. Helleberg M, Niemann CU, Moestrup KS, et al. Persistent COVID-19 in an Immunocompromised Patient Temporarily Responsive to Two Courses of Remdesivir Therapy. J Infect Dis 2020; 222(7): 1103–7.

41. Sepulcri C, Dentone C, Mikulska M, et al. The longest persistence of viable SARS-CoV-2 with recurrence of viremia and relapsing symptomatic COVID-19 in an immunocompromised patient – a case study. medRxiv 2021: 2021.01.23.21249554.

42. McMahan K, Yu J, Mercado NB, et al. Correlates of protection against SARS-CoV-2 in rhesus macaques. Nature 2020.

43. Rausch JW, Capoferri AA, Katusiime MG, Patro SC, Kearney MF. Low genetic diversity may be an Achilles heel of SARS-CoV-2. Proc Natl Acad Sci U S A 2020; 117(40): 24614–6.

