## Supplemental doc for "SARS-CoV-2 Sequence Characteristics of COVID-19 Persistence and Reinfection"

**Supplemental Figure 1.** (A) PRISMA flowchart for reinfection literature search. (B) PRISMA flowchart for persistent infection literature search.

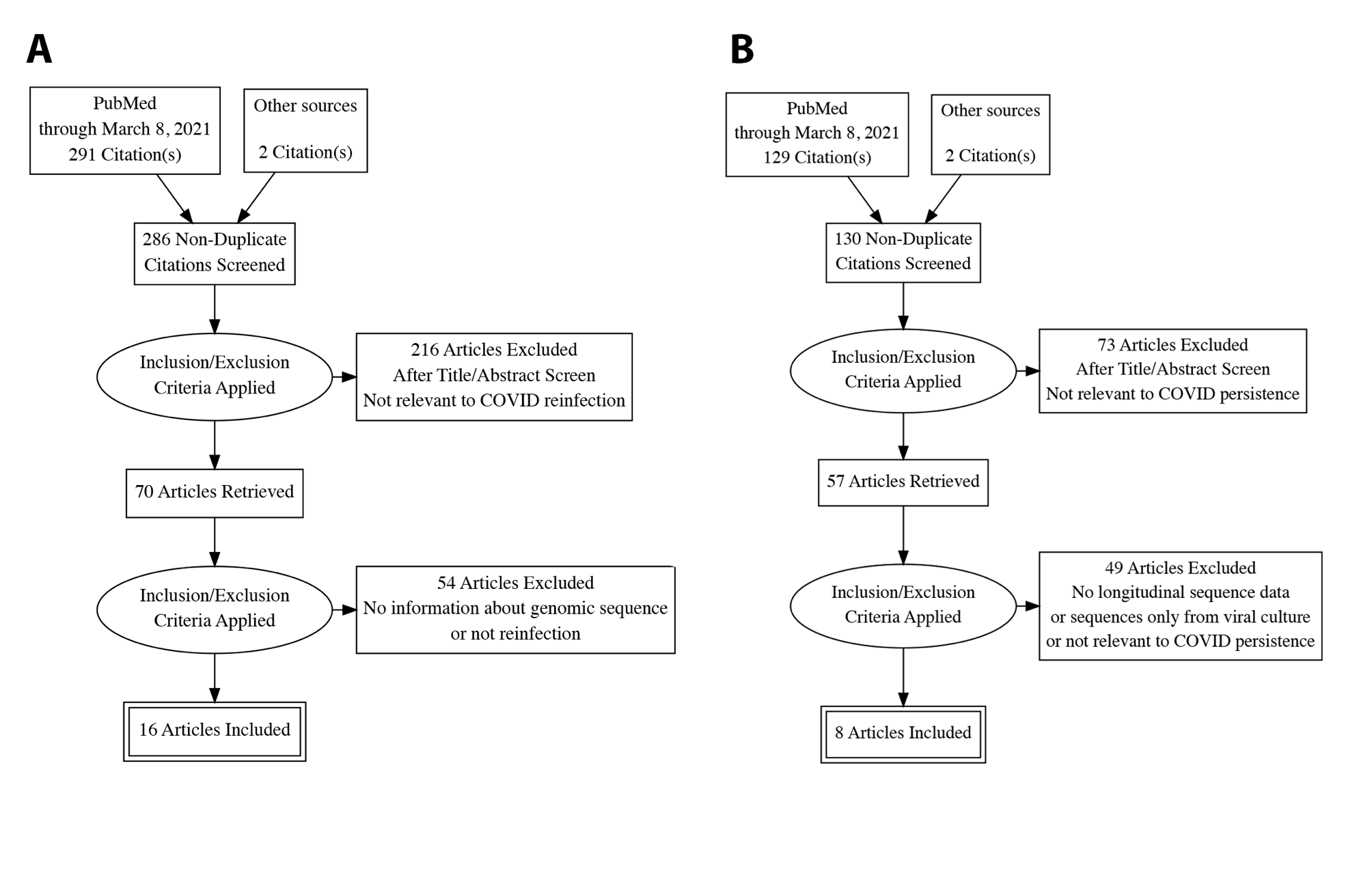

**Supplemental Figure 2.** Maximum likelihood phylogenetic tree as shown in Figure 1, but with unclassified cases highlighted.

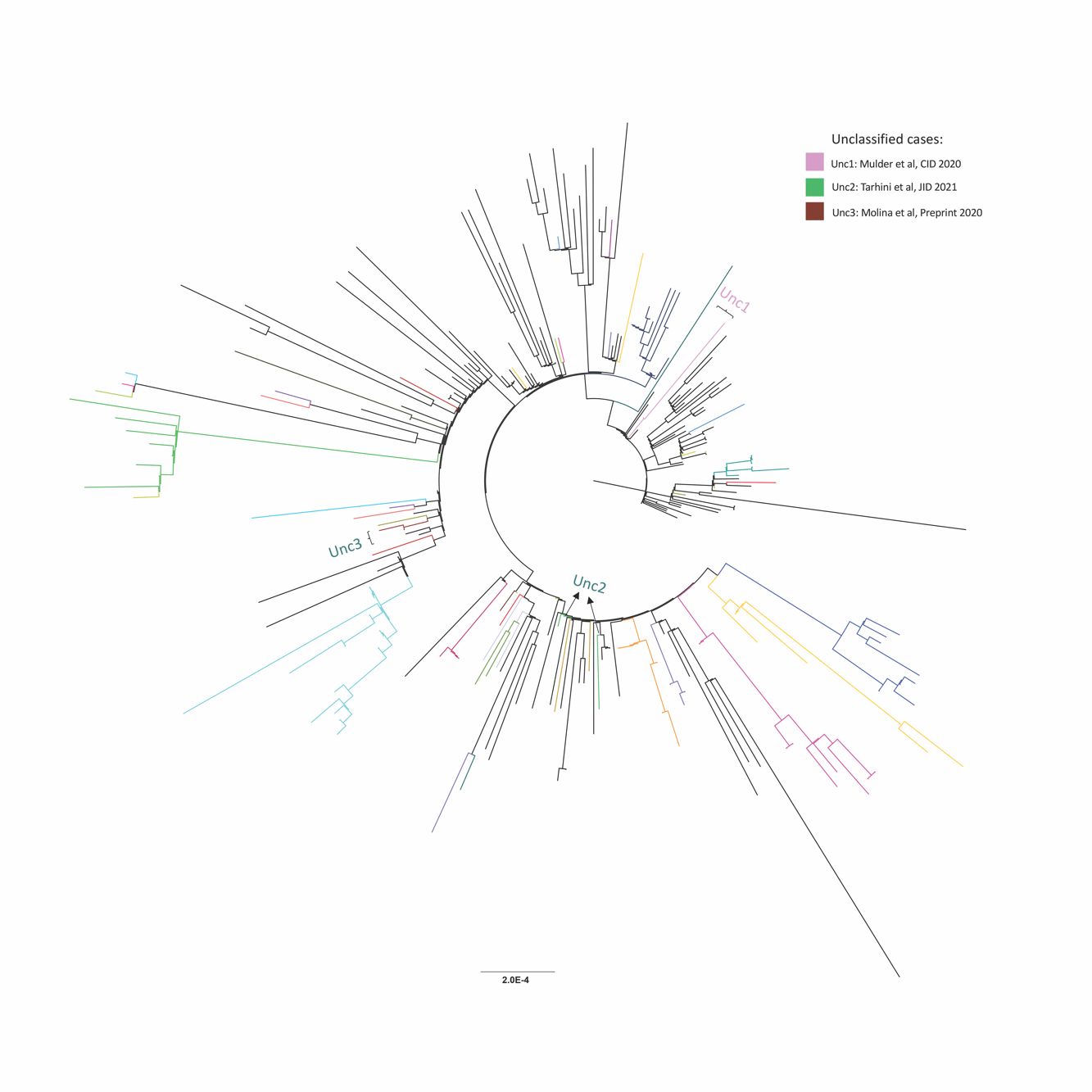

**Supplemental Figure 3.** (A) Amino acid substitution frequency pooled across all reinfection cases for each SARS-CoV-2 gene. Dashed line indicates global substitution frequency across the whole genome. Substitution frequency for each gene was compared to the substitution frequency in the rest of the genome using a Fisher’s exact test. P-values were corrected for multiple comparisons using the Bonferroni correction. * <0.05, ** <0.01 and ***<0.001. (B) Nucleotide changes in the second infection relative to the first infection by clinical disease severity. Mutations shown for the whole genome and S gene. P=0.67, Mann Whitney test. (C) Number of rare amino acid changes at each time point relative to circulating sequences in the community. P=0.77, Wilcoxon matched-pairs signed rank test. ORF: open reading frame, S: Spike, E: Envelope, M: Membrane, N: Nucleocapsid.

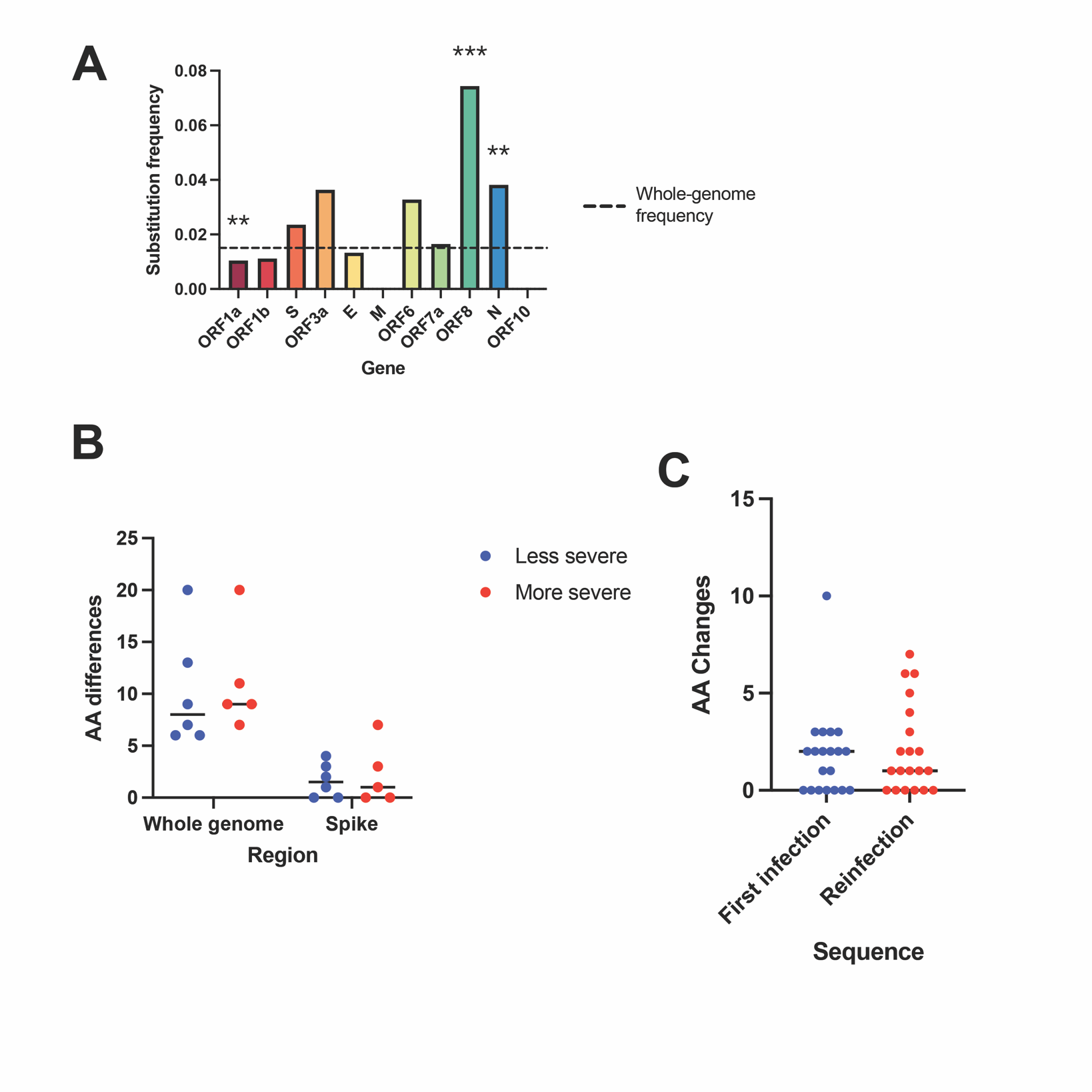

**Supplemental Figure 4.** Circos plots mapping mutations for the persistent COVID-19 cases. Mutations are marked relative to the first timepoint. Pe1 * shows N501Y mutation (days 128-152). ** shows E484K mutation (days 75 and 81 only). Pe4 *** shows Δ69/Δ70 mutation. Pe6-3 **** shows N440K and E484Q. Several patients Δ shows deletions of some or all of Spike residues 141-145. Red text indicates timepoint was sampled after first convalescent plasma or antibody cocktail treatment. Inner ticks indicate nucleotide position. ORF: open reading frame, S: Spike, E: Envelope, M: Membrane, N: Nucleocapsid.

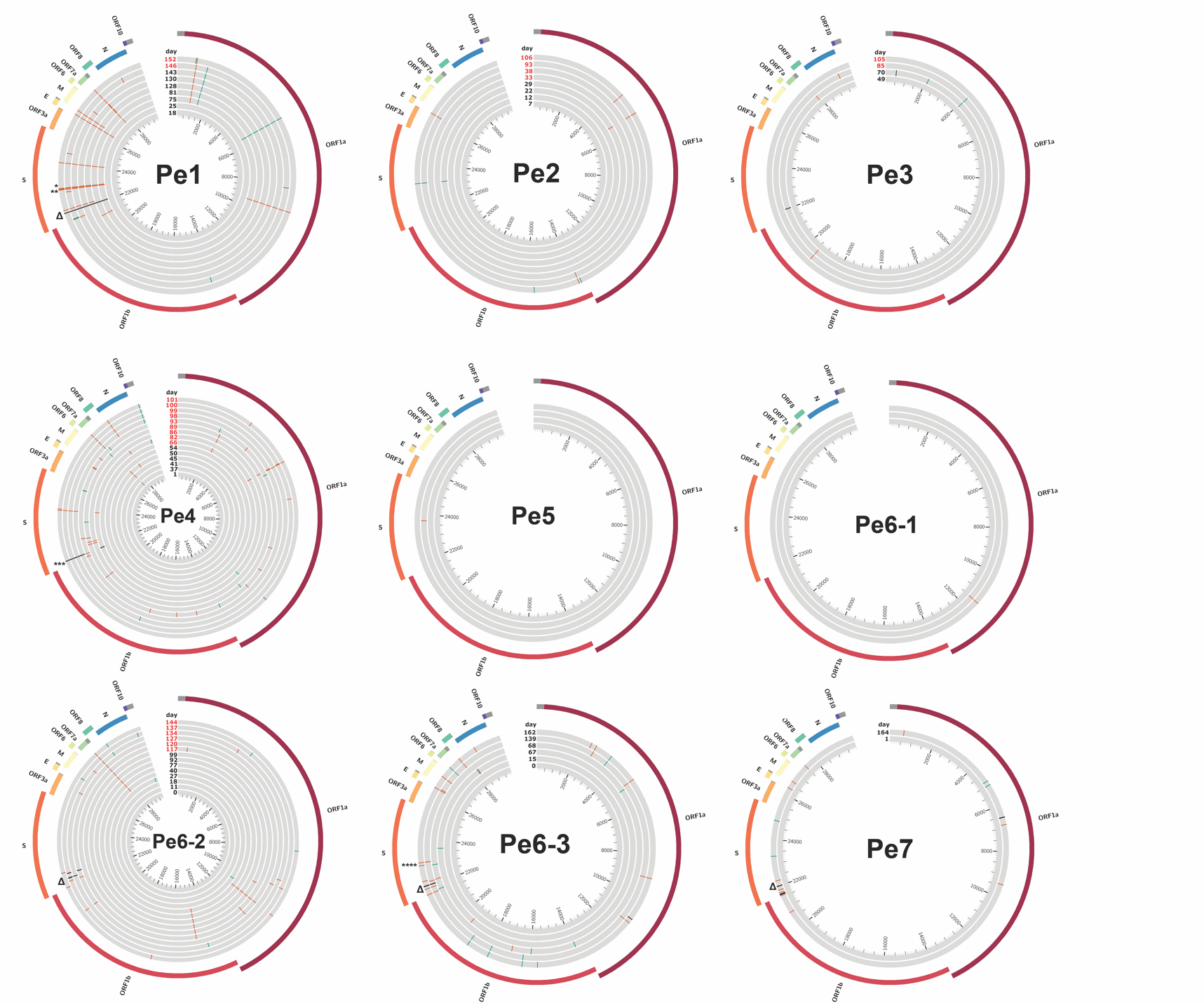

**Supplemental Figure 5.** Accumulation of nonsynonymous changes in persistent COVID-19 cases. (A) Amino acid (AA) substitution frequency pooled across all persistent cases for each SARS-CoV-2 gene. Dashed line indicates global substitution frequency across the whole genome. Substitution frequency for each gene was compared to the substitution frequency in the rest of the genome using a Fisher’s exact test. P-values were corrected for multiple comparisons using the Bonferroni correction. * <0.05, ** <0.01 and ***<0.001 (B) Amino acid changes in samples taken prior to convalescent plasma or monoclonal antibody treatment relative to first sampled sequence in each persistently infected patient. Regression line and 95% confidence bands are shown. (C) Amino acid changes in samples taken after convalescent plasma or monoclonal antibody treatment relative to last sample taken prior to treatment in each persistently infected patient. Regression line and 95% confidence bands are shown.

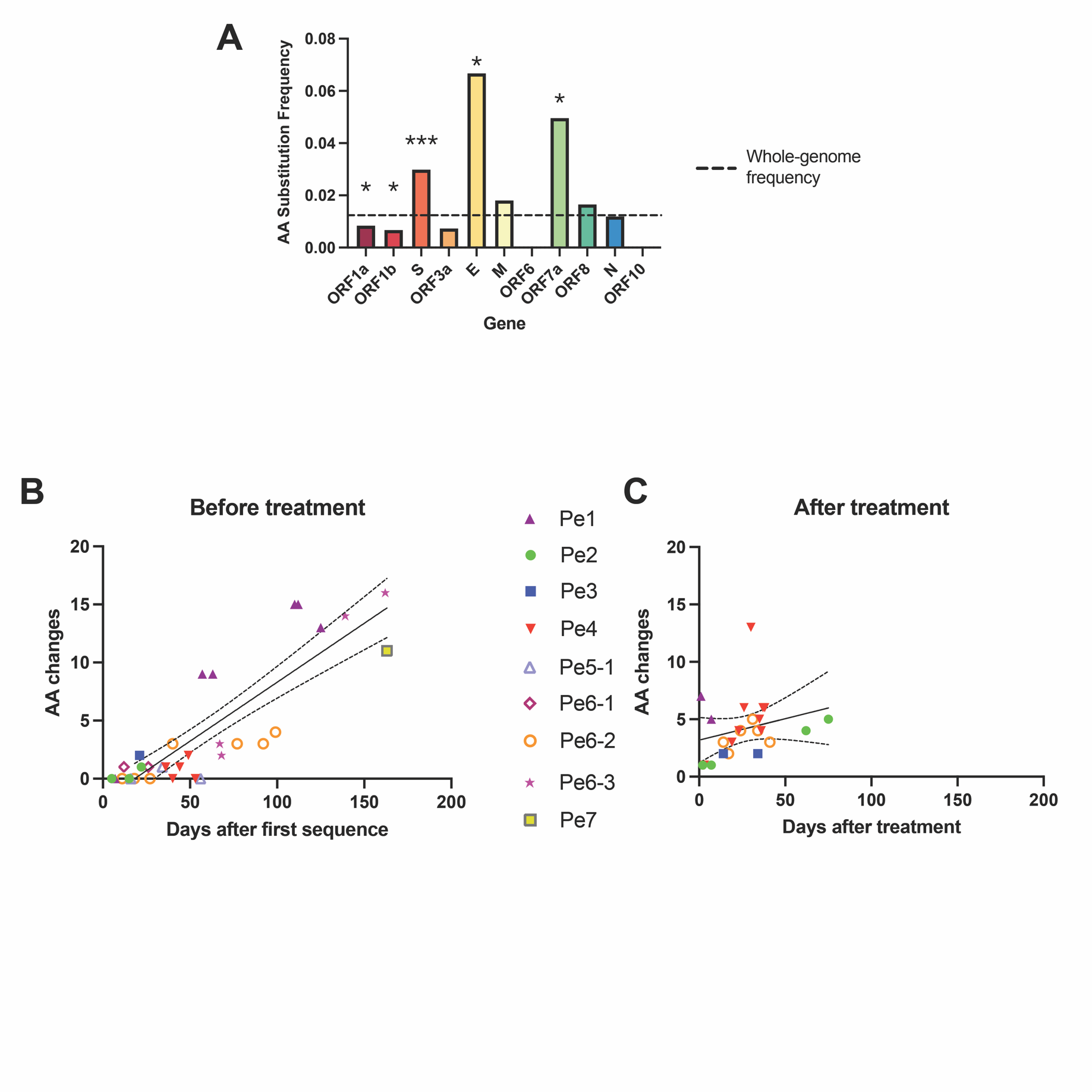

**Supplemental Table 1.** Unclassified cases

| Patient | Case | Publication (year) | Age | Sex | Underlying conditions | Immunosupressants | Time between sequences | Reason for lack of classification |
| --- | --- | --- | --- | --- | --- | --- | --- | --- |
| Unc 1 | Mulder, *et al.* | *Clin. Infect. Dis.* (2020) | 89 | F | Waldenström macroglobulinemia | B cell-depleting therapy | 59 | Originally reported as reinfection; genomic data points to persistence, but no intervening sampling to confirm |
| Unc 2 | Tarhini, *et al.* | *J. Infect. Dis.* (2021) | 35 | M | Rheumatoid arthritis | Rituximab | 82 | 3^rd^ sequence does not branch with first 2, coinfection hypothesisi |
| Unc 3 | Molina, *et al.* | Research Square (2020) | 35 | M | None | None | 181 | Patient characteristic not consistent with other persistent cases, number of mutations over timeframe inconsistent with persistence or reinfection |

**Supplemental Table 2. Information on sequences used to compare reinfection sequences to circulating virus strains.** Sequences from the indicated geographic location, clade, and within one month of each infection timepoint were downloaded from GISAID (see Supplemental Methods and the accession numbers are listed in Supplemental Data 2.

| **Case** | **Timepoint** | **Date** | **Clade** | **Location** | **Locations sampled** | **Sequences retrieved** |
| --- | --- | --- | --- | --- | --- | --- |
| Re1 | First | March 2020 | 19A | Belgium | Belgium | 41 |
|  | Second | September 2020 | 20A |  |  | 119 |
| Re2 | First | March 2020 | 20A | Hong Kong | Hong Kong | 17 |
|  | Second | August 2020 | 20E | Spain or United Kingdom | Spain, United Kingdom | 2010 |
| Re3 | First | May 2020 | 20A | Ecuador | Ecuador, Peru, Colombia | 43 |
|  | Second | July 2020 | 19B |  | Global | 195 |
| Re4 | First | April 2020 | 20C | Nevada, U.S. | Nevada, U.S. | 92 |
|  | Second | June 2020 | 20C |  |  | 31 |
| Re5 | First | March 2020 | 19B | Washington state, U.S. | Washington state, U.S. | 1063 |
|  | Second | July 2020 | 20A |  |  | 232 |
| Re6 | First | June 2020 | 20B | Brazil | Brazil | 228 |
|  | Second | October 2020 | 20B |  |  | 149 |
| Re7 | First | April 2020 | 19A | United Kingdom | United Kingdom | 751 |
|  | Second | December 2020 | 501Y.V1 |  |  | 3991 |
| Re8 | First | March 2020 | 20B | Belgium | Belgium | 271 |
|  | Second | June 2020 | 19B |  | Europe (except United Kingdom) | 68 |
| Re9 | First | April 2020 | 20A | France | France | 415 |
|  | Second | August 2020 | 20A.EU2 |  |  | 111 |
| Re10 | First | May 2020 | 20B | Brazil | Brazil | 431 |
|  | Second | October 2020 | 20B |  |  | 150 |
| Re11-1 | First | May 2020 | 19A | India | Global | 369 |
|  | Second | August 2020 | 20A |  | India | 189 |
| Re11-2 | First | May 2020 | 20A | India | India | 491 |
|  | Second | September 2020 | 20A |  |  | 189 |
| Re12-1 and Re12-2 | First | April 2020 | 19A | Qatar | Qatar, Oman, Israel, United Arab Emirates, Jordan, Georgia, Saudi Arabia | 27 |
|  | Second | June/July 2020 | 20A |  |  | 93 |
| Re13-1 | First | March 2020 | 20A | Brazil | Brazil | 141 |
|  | Second | December 2020 | 20J/501Y.V3 |  |  | 426 |
| Re13-2 | First | October 2020 | 20B | Brazil | Brazil | 317 |
|  | Second | January 2021 | 20J/501Y.V3 |  |  | 426 |
| Re13-3 | First | April 2020 | 20A | Brazil | Brazil | 172 |
|  | Second | January 2021 | 20J/501Y.V3 |  |  | 426 |
| Re14 | First | April 2020 | 20A | Switzerland | Switzerland | 790 |
|  | Second | October 2020 | 20A.EU2 |  |  | 1430 |
| Re15 | First | August 2020 | 20B | India | India | 764 |
|  | Second | November 2020 | 20B |  |  | 43 |
| Re16 | First | May 2020 | 20A | Brazil | Brazil | 58 |
|  | Second | July 2020 | 20A |  |  | 35 |
| Unc1* | First | April 2020 | 19A | The Netherlands | The Netherlands | 36 |
|  | Second | June 2020 | 19A |  | Europe | 58 |

*Although we did not classify this case, we still compared the sequences to circulating sequences to rule out reinfection.

**Supplemental Table 3.** Spike mutations observed in reinfection cases

| **First sequence** | **Second sequence** | **Proportion of cases** |
| --- | --- | --- |
| D614 | D614G | 8/20 |
| E484/V1176 | E484K/V1176F | 2/20 |
| N440 | N440K (escape variant) | 1/20 |
|  | del22832 | 1/20 |
| K790 | K790I | 1/20 |
| V289/S477/D578 | V289E/S477N/D578A | 1/20 |
| S477 | S477N | 1/20 |
| K417/E484/N501 | K417T/E484K/N501Y | 3/20 |

**Supplemental Table 4.** Mean, Median, and 95% HPD interval for the substitution rates of global sequences and each of the persistent patients.

|  | **Mean** | **Median** | **95% HPD* Interval** |
| --- | --- | --- | --- |
| Global | 6.77E-04 | 6.76E-04 | [5.91E-4, 7.66E-4] |
| Pe1 | 5.29E-03 | 4.42E-03 | [3.26E-4, 0.0125] |
| Pe2 | 1.59E-03 | 1.19E-03 | [8.31E-5, 4.18E-3] |
| Pe3 | 3.42E-03 | 2.62E-03 | [1.33E-4, 8.84E-3] |
| Pe4 | 3.32E-03 | 2.88E-03 | [2.91E-4, 7.42E-3] |
| Pe5-1 | 1.80E-03 | 1.09E-03 | [4.01E-6, 5.75E-3] |
| Pe6-1 | 4.48E-03 | 2.94E-03 | [4.60E-5, 0.0139] |
| Pe6-2 | 2.96E-03 | 2.52E-03 | [1.99E-4, 6.67E-3] |
| Pe6-3 | 6.67E-03 | 5.30E-03 | [5.27E-4, 0.0164] |

*95% HPD: the 95% Highest Posterior Density of the substitution rates.

**Supplemental Methods**

**Unclassified cases**

Several cases were not included in this analysis due to uncertainty in classification (Supplemental Table 1; Supplemental Figure 2). One, reported in Mulder, *et al.* as a case of reinfection, was an elderly patient with Waldenström macroglobulinemia treated with B cell-depleting therapy (Unc1) [1]. This case shared features persistent infection, including immunosuppression and sequences that clustered together in the phylogenetic analysis. However, there was some uncertainty given that only two sequences were available. Another, Unc2, was described as a persistent patient treated with B cell-depleting therapy, with two sequences clustering together but a third sequence clustering independently. The authors proposed this to be a possible case of both persistence and coinfection [2]. Another case (Unc3), involving a healthy individual with two episodes separated by 160 days, was an outlier with a low number of mutations between sequences and there was uncertainty about whether this case represented reinfection or persistent infection [3].

**Phylogenetic tree sampling**

SARS-CoV-2 sequences were sampled from GISAID to be globally and temporally representative of sequence diversity. Large samples of the variants of concern B.1.1.7 and B.1.351 were also included. GISAID accession numbers are included in Supplemental Data 1.

**Reinfection sequence comparison**

For the first and second sequence of each reinfection case, contemporaneous viral sequences circulating in the same geographic region, within one month, and from the same NextStrain clade were downloaded from GISAID to identify rare mutations in the reinfection sequences. The geographic location was sometimes broader than the location the infection occurred so that a larger number of sequences are represented. GISAID accession numbers for each patient’s sequences are listed in Supplemental Data 2. Information about clade, location, and time frame for each patient is listed in Supplemental Table 2.

**Supplemental References**

1.	Mulder M, van der Vegt D, Oude Munnink BB, et al. Reinfection of SARS-CoV-2 in an immunocompromised patient: a case report. Clin Infect Dis **2020**.

2.	Tarhini H, Recoing A, Bridier-Nahmias A, et al. Long term SARS-CoV-2 infectiousness among three immunocompromised patients: from prolonged viral shedding to SARS-CoV-2 superinfection. J Infect Dis **2021**.

3.	Molina LP, Chow SK, Nickel A, Love JE. Prolonged Detection of Severe Acute Respiratory Syndrome Coronavirus 2 (SARS-CoV-2) RNA in an Obstetric Patient With Antibody Seroconversion. Obstet Gynecol **2020**.
